# Evaluation of Disk Potentiation Test (DPT) and Double Disk Synergy Test (DDST) for The Detection of Metallo-β-lactamases (MBLs) in Clinical Isolates of Bangladesh

**DOI:** 10.1101/2022.12.24.22283822

**Authors:** Sumon Kumar Das, Afzal Sheikh, Nikhat Ara, Suma Mita Biswas, Abhinandan Chowdhury, Fatimah Az Zahra, Chaman Ara Keya

## Abstract

**Objective:** Increasing the emergence of Metallo-β-lactamase (MBL) producing gram-negative *Enterobacteriaceae* and their dexterous horizontal transmission of the gene among other strains, demands rapid and accurate detection. This study was conducted to determine a suitable MBL detection method that could promptly identify the distribution of MBL-producing Gram-negative isolates at hospital settings in Bangladesh.

**Methods:** A total of 103 gram-negative bacilli were identified from various clinical samples at a tertiary care hospital in Dhaka city. MBL producers were detected by two phenotypic methods; Disk Potentiation Test (DPT) and the Double Disk Synergy Test (DDST) based on β-lactam chelator combinations where EDTA/SMA has been used as inhibitor and Imipenem, Ceftazidime as substrates.

**Results:** All 103 isolates which were identified as *Escherichia coli spp, Klebsiella spp, Pseudomonas spp, Acinetobacter spp, Proteus spp, Providencia spp* were found to be multidrug-resistant in antibiogram test. All the mentioned isolates showed complete resistance (100%) to Imipenem, Meropenem, and Amoxiclav. The highest carbapenem-resistant etiological agents isolated were *Acinetobacter spp* 40 (38.8%) followed by *Pseudomonas spp* 27 (26.2%), *Klebsiella spp* 26 (25.2%), *Escherichia coli* 8(7.8%), *Proteus spp* 1(1%) and *Providencia spp* 1(1%). DPT method detected significantly (p=0.000009) higher number of MBL-producers (n=61, 59.2% & n=56, 54.4%) compared to the DDST method (n=43, 41.7%, n=38, 36.9% & n=15, 14.6%).

**Conclusion:** This study depicts that DPT is a more sensitive method than DDST and could be recommended for identifying MBL-producing bacteria in Bangladeshi hospitals for the proper management of patients, and to reduce time constraints as well as treatment costs.

## Introduction

Carbapenemases including Metallo-β-lactamases (MBLs) producing bacteria (*Enterobacteriaceae*) are of paramount global concern given limited therapeutic options, and untoward clinical outcomes. The horizontal transmission of carbapenemase genes mediated by mobile genetic elements (self-transmissible plasmids) carrying additional resistance elements which confer resistance to various groups of antibiotics, results in multidrug resistance, including bacteria resistant to all available antibiotics (1-2). Due to their marked resistance to a wide range of antibiotics, infections caused by carbapenemase-producing *Enterobacteriaceae* are terribly difficult to manage. Moreover, *Enterobacteriaceae*, being a part of intestinal flora are easily spread and difficult to eliminate, especially countries like Bangladesh with low levels of hygiene. The widespread use of antibiotics in humans and in the food chain and their spillover into the environment accelerate the development, selection, and/or horizontal transfer of antibiotic resistance plasmids in a given bacterial population (3-4). Since MBLs are mostly transposon-and/or integron-encoded determinants that can easily disseminate to other enterobacterial strains (5-8).

It is conceivable that the primary source of these bacteria are hospitals and other healthcare settings where severe cases of bacterial infections are presented, and the volume of antibiotic use is high (9-10). Numerous studies have described and evaluated the performance of simple phenotypic tests for the specific detection of carbapenemase-producing strains by different groups of scientists all over the globe, with a marked endemicity according to enzyme type (1). It appears that the effect of Carbapenem-resistant *Enterobacteriaceae* (CRE) in Bangladesh and their impact in the environment are uncharted. However, to our knowledge, only one study for phenotypic detection of MBLs by DPT has been done in Bangladesh (11). However, none of the researchers performed comparative studies among the MBL detection methods.

Therefore, practical and accurate phenotypic approaches are urgently needed to detect obviously increasing carbapenemase producers among *Enterobacteriaceae* in the clinical laboratory or diagnostic laboratories lacking molecular identification setup. These assays may provide substantial information before application of the more expensive and sophisticated molecular techniques(5,12). Furthermore, phenotypic methods will include all novel carbapenemase gene types (1) which ‘the predefined gene target’ based PCR method cannot identify.

Considering the public health threat, the MBL producers pose and the rapid dissemination of MBL in bacteria, the systemic search for MBL detection methods based on β-lactam-chelator combinations seems praiseworthy which could perform well to identify all MBL producing enterobacterial species in Bangladesh. Therefore, the present study was aimed to determine an accurate phenotypic method through comparative studies among Disc Potentiation Test (DPT) & Double Disc Synergy Test (DDST) to explore the incidence of multidrug-resistant MBL producers in Bangladesh along with antimicrobial-resistant patterns of these organisms.

## Methods

### Sample collection

A total of 103 CRE isolated from various unit of Evercare hospital, previously called Apollo Hospital, Dhaka, were used for this study. These included (i) 13 isolates from outpatient (OPD) (ii) 43 isolates from inpatients (IPD) (iii) 47 isolates from Intensive care unit (ICU). The ratio of infected male to female, as well as infection rate in various age-groups were recorded accordingly. Bacterial samples were isolated from different clinical specimen including Urine, Tracheal aspirate, Sputum, Blood, ET tube tips, Pus, Suction tips, Wound swab, Foleys catheter tips, Silicon catheter, CVP tips. Oral consent was obtained from the patients and the data were analyzed anonymously. This study was reviewed and approved by “Research and Ethical Practice Committee” of Evercare Hospital, Dhaka, Bangladesh.

### Antimicrobial Susceptibility Testing

All these specimens were inoculated on appropriate culture media (MacConkey’s agar, Blood agar, Chocolate agar and only for urine sample we used HiCrome UTI agar) and incubated for 24 to 48 hours at 37[C. After incubation organisms were identified by standard microbiological procedures (Colonial morphology, Gram’s stain appearance, catalase test, oxidase test, triple sugar iron test, motility test, urease test, citrate test and indole test). Antibiotic susceptibility tests were performed using the Kirby-Bauer disc diffusion techniques according to CLSI guidelines (23). Inoculates were prepared by suspending the isolates in normal saline equal to turbidity of 0.5 McFarland turbidity standard and applied on muller Hinton Agar plates. All antibiotic discs were obtained from Oxoid, UK. Antibiotic discs were used depending on the types of microorganism and specimens (Amoxiclav 30 mcg, Ceftriaxone 30 mcg, Cefepime 30 mcg, Ceftazidime 30 mcg, Imipenem 10 mcg, Meropenem 10 mcg, Polymyxin 300 mcg, Amikacin 30 mcg, Gentamicin 10 mcg, Netilmicin 30 mcg, Tetracycline 30 mcg, Tigecycline 15 mcg, Ciprofloxacin 5 mcg, Levofloxacin 5 mcg, Cotrimoxazole 25 mcg). These discs were incubated along with controls for 18-24 hours at 37□C aerobically.

Isolated Gram-negative bacteria were subjected to an array of antibiotics and the results were interpreted as resistant or sensitive according to criteria set by Clinical and Laboratory Standards Institute (CLSI). Gram-negative isolates with Imipenem (IPM) resistance were tested for detection of MBL by disc potentiation test (DPT) and double disc synergy test (DDST) method. A total number of one hundred three Gram-negative bacteria showed Carbapenem resistance in routine culture.

### Double Disc Synergy Test (DDST)

The bacterial isolates were sub-cultured on Mac-Conkey agar media and incubated overnight at 37 °C. After overnight incubation, 2-3 isolated colonies of the organism were picked up from the subculture plate by a sterile inoculating wire loop and suspended in 3ml sterile peptone water in a screw-capped test tube. Test strain was adjusted to the McFarland 0.5 standard. Within 15 minutes of adjustment of density of the organism, streaking on Mueller Hinton agar was done using a sterile cotton swab.

One disc containing Ceftazidime (CAZ) (30 mcg) and two IPM (10 mcg) were placed on the plate. The distance between every CAZ/IPM disc was kept at about 4 cm from center to center (13). Two blank discs with 10 μl of 0.1 M EDTA added was placed near the CAZ/IPM disc, within a center-to-center distance 2 cm. A Sodium mercaptoacetate (SMA) (Metallo-β-lactamase SMA Eiken; Eiken Chemical Co., Ltd., Tokyo, Japan) disc was placed near the IPM disc respectively disc distance edge to edge 10 mm. The agar plate was incubated at 37 °C overnight (14,15).

In Double Disc Synergy Test (DDST) with 0.1 M EDTA, the enhancement of synergistic growth inhibition zones between the CAZ/IPM disc and disc containing 0.1M EDTA was considered as positive for MBLs (13). The presence of synergistic inhibition zone of IPM with greater than 5 mm enlargement with the SMA disc side, was interpreted as positive (15).

### Disc Potentiation Test (DPT)

The bacterial isolates were sub-cultured on Mac-Conkey agar media and incubated overnight at 37 °C. After overnight incubation; 2-3 isolated colonies of the organism were suspended in 3ml sterile peptone water. The test strain was adjusted to the McFarland 0.5 standard. The bacterial suspension was later streaked on Mueller Hinton agar media.

Disc Potentiation Test (DPT) was done as per Yong et al using IPM-EDTA (16,17) The test depends on comparing the zones given by disc containing IPM with or without EDTA modified the test by using two 10 mcg IPM disc and two 30 mcg Ceftazidime (CAZ) disc (17,18). A zone diameter difference between the IPM/CAZ and IPM/CAZ+ EDTA disc >7 mm was interpreted as a positive result.

Two discs containing 30 mcg CAZ and two discs containing 10 mcg IPM were placed on the plates. The distance between every CAZ/IPM disc was kept at about 4 cm from center to center. 10 μl of 0.1 M EDTA was added to one CAZ/IPM disc. The plate was incubated at 37°C overnight.

In this study DPT with 0.1 M EDTA, the enlargement of the diameter of growth inhibitory zone around CAZ/IPM + EDTA disc with greater than 7 mm, compared to CAZ/IPM alone was considered as positive for MBLs.

Statistical analysis: For the statistical analyses, data were entered into an excel spreadsheet and analyzed using the statistical software SPSS version 20.0 (IBM; Chicago, IL, USA) for Windows was utilized. Bivariate correlation test. A p value of <0.05 was considered as statistically significant.

## Results

### Bacterial isolates from all age-groups showed 100% resistance to Amoxiclav, and Carbapenems while “61-75” age-group showed maximum number of resistant isolates

A total of 103 gram-negative strains were isolated from different clinical specimens. All these bacteria isolated from the patients, were subjected to antibiogram test. Antibiotic susceptibility profile (n=103) was analyzed for the seven isolated bacterial spps. All 103 isolates showed 100% resistance to Imipenem, Meropenem, and Amoxiclav. We also investigated the distribution of resistant and susceptible isolates among male and female individuals. Results represented that the number of resistant isolates distribution was significantly high in male patients compare to the females (Table 1.). The percentage of bacterial isolates from male and female patient groups that showed 100% resistant to Imipenem, Meropenem, and Amoxiclav were 70% and 30 % respectively (Figure 1.a). We also observed that *Escherichia coli spp, Klebsiella spp, Pseudomonas spp, Acinetobacter spp, Proteus spp*, and *Providencia spp*. showed a significantly higher distribution among male patient group compare to female patient group (Figure 1.b). In addition, our observation of the distribution resistant bacterial isolates (n=103) among age-groups data revealed that “61-75” age-group showed the maximum number of resistant isolates. The age-group also represented the maximum resistant isolates from ICU hospital unit compare to OPD, IPD units (Figure 1c). The levels of significance that has been analyzed statistically by Pearson-correlation has been shown in supplement Table 1. We investigated if particular samples type is useful for testing susceptibility of clinically isolated etiological agents with antibiotics for proper managing of the patients. We analyzed sample wise distribution of bacterial isolates, and found that Tracheal aspirate, followed by Urine and Wound swab samples represented maximum number of resistant isolates (Figure 2., and Table 2.). While all the bacteria isolated were found to be resistant to the antibiotic arsenal, the highest carbapenem-resistant etiological agents isolated was *Acinetobacter spp* 40 (38.8%) followed by *Pseudomonas spp* 27 (26.2%), *Klebsiella spp* 26 (25.2%), *Escherichia coli* 8 (7.8%), *Proteus spp* 1 (1%) and *Providencia spp* 1 (1%) (Table 3.). Our investigation on the susceptibility of bacterial isolates (n=103) from all age-groups to antibiotic arsenal, (Amoxiclav, Ceftriaxone, Cefepime, Ceftazidime, Imipenem, Meropenem, Polymyxin, Amikacin, Gentamicin, Netilmicin, Tetracycline, Tigecycline, Ciprofloxacin, Levofloxacin, and Cotrimoxazole) showed that all the bacterial isolates showed 100% resistance to Amoxiclav (30 mcg) and carbapenem (imipenem (10 mcg), and Meropenem (10 mcg) (Table 4.).

**Table 1.** Antibiotic susceptibility profiles of different Carbapenem-resistant Enterobacteriaceae (CRE, n=103). All 103 isolates showed 100% resistance to Imipenem, Meropenem, and Amoxiclav. The antibiotic susceptibility profile was analyzed for *Escherichia coli spp, Klebsiella spp, Pseudomonas spp, Acinetobacter spp, Proteus spp, Providencia spp* (n=103) against antibiotics using the Kirby-Bauer disc diffusion techniques according to CLSI guidelines. The distribution of resistant and susceptible isolates among males and females was also analyzed. Results represented that the number of resistant isolates distribution was significantly high in male patients compare to the females

|  |  |  | Gender |  |  |  | Total |  |
| --- | --- | --- | --- | --- | --- | --- | --- | --- |
|  |  |  | Male |  | Female |  |  |  |
| Modes of Action | Antimicrobial Category | Antibiotics | Sensitive | Resistant | Sensitive | Resistant | Sensitive | Resistant |
| Cell wall synthesis inhibitors | Aminopenicillin | Amoxiclav (30 mcg) | - | 72 | - | 31 | - | 103 |
|  | Extended spectrum Cephalosporins | Ceftriaxone (30 mcg) | - | 72 | 1 | 30 | 1 | 102 |
|  |  | Cefepime (30 mcg) | 3 | 69 | 1 | 30 | 4 | 99 |
|  |  | Ceftazidime (30 mcg) | 3 | 69 | 2 | 29 | 5 | 98 |
|  | Carbapenems | Imipenem (10 mcg) | - | 72 | - | 31 | - | 103 |
|  |  | Meropenem (10 mcg) | - | 72 | - | 31 | - | 103 |
| Cell membrane | Polypeptide antibiotics | Polymyxin (300 mcg) | 63 | 9 | 31 | - | 94 | 9 |
| Protein synthesis inhibitors | Aminoglycosides | Amikacin (30 mcg) | 8 | 64 | 3 | 28 | 11 | 92 |
|  |  | Gentamicin (10 mcg) | 7 | 65 | 4 | 27 | 11 | 92 |
|  |  | Netilmicin (30 mcg) | 6 | 66 | 3 | 28 | 9 | 94 |
|  | Tetracyclines | Tetracycline (30 mcg) | 12 | 60 | 7 | 24 | 19 | 84 |
|  |  | Tigecycline (15 mcg) | 55 | 17 | 28 | 3 | 83 | 20 |
| Nucleic acid synthesis inhibitors | Fluoroquinolones | Ciprofloxacin (5 mcg) | 2 | 70 | 2 | 29 | 4 | 99 |
|  |  | Levofloxacin (5 mcg) | 4 | 68 | 2 | 29 | 6 | 97 |
| Folate pathway inhibitors | Trimethoprim and Sulfamethoxazole | Cotrimoxazole (25 mcg) | 9 | 63 | 5 | 26 | 14 | 89 |

**Table 2.** Sample-wise distribution spectrum of various (CRE) bacterial isolates. individual bacterial isolates (n=103) from Urine, Tracheal aspirate, Sputum, Blood, ET tube tips, Pus, Suction tips, Wound swab, Foleys catheter tips, Silicon catheter, and CVP tips were treated against commonly used antimicrobial agents. Escherichia coli spp, Klebsiella spp, Pseudomonas spp, Acinetobacter spp, Proteus spp, and Providencia spp showed resistant to antimicrobial agents.

| Antimicrobial agents | Clinical samples |  |  |  |  |  |  |  |  |  |  |  |  |  |  |  |  |  |  |  |  |  |
| --- | --- | --- | --- | --- | --- | --- | --- | --- | --- | --- | --- | --- | --- | --- | --- | --- | --- | --- | --- | --- | --- | --- |
|  | Urine |  | Tracheal aspirate |  | Sputum |  | Blood |  | ET tube tips |  | Pus |  | Suction tips |  | Wound swab |  | Foleys catheter tips |  | Silicon catheter |  | CVP tips |  |
|  | S | R | S | R | S | R | S | R | S | R | S | R | S | R | S | R | S | R | S | R | S | R |
| Amoxiclav (30 mcg) | - | 37 | - | 28 | - | 6 | - | 4 | - | 5 | - | 4 | - | 2 | - | 12 | - | 3 | - | 1 | - | 1 |
| Ceftriaxone (30 mcg) | - | 37 | - | 28 | - | 6 | - | 4 | 1 | 4 | - | 4 | - | 2 | - | 12 | - | 3 | - | 1 | - | 1 |
| Cefepime (30 mcg) | 1 | 36 | 3 | 25 | - | 6 | - | 4 | - | 5 | - | 4 | - | 2 | - | 12 | - | 3 | - | 1 | - | 1 |
| Ceftazidime (30 mcg) | 1 | 36 | 3 | 25 | - | 6 | - | 4 | 1 | 4 | - | 4 | - | 2 | - | 12 | - | 3 | - | 1 | - | 1 |
| Imipenem (10 mcg) | - | 37 | - | 28 | - | 6 | - | 4 | - | 5 | - | 4 | - | 2 | - | 12 | - | 3 | - | 1 | - | 1 |
| Meropenem (10 mcg) | - | 37 | - | 28 | - | 6 | - | 4 | - | 5 | - | 4 | - | 2 | - | 12 | - | 3 | - | 1 | - | 1 |
| Polymyxin (300 mcg) | 31 | 6 | 26 | 2 | 6 | - | 4 | - | 5 | - | 4 | - | 2 | - | 12 | - | 3 | - | - | 1 | 1 | - |
| Amikacin (30 mcg) | 4 | 33 | 2 | 26 | - | 6 | 1 | 3 | - | 5 | 2 | 2 | - | 2 | 1 | 11 | 1 | 2 | - | 1 | - | 1 |
| Gentamicin (10 mcg) | 6 | 31 | 1 | 27 | - | 6 | 1 | 3 | 1 | 4 | 1 | 3 | - | 2 | - | 12 | 1 | 2 | - | 1 | - | 1 |
| Netilmicin (30 mcg) | 2 | 35 | 1 | 27 | - | 6 | 1 | 3 | 1 | 4 | 2 | 2 | 1 | 1 | - | 12 | 1 | 2 | - | 1 | - | 1 |
| Tetracycline (30 mcg) | 8 | 29 | 5 | 23 | 1 | 5 | 1 | 3 | 2 | 3 | - | 4 | - | 2 | 2 | 10 | - | 3 | - | 1 | - | 1 |
| Tigecycline (15 mcg) | 28 | 9 | 22 | 6 | 4 | 2 | 4 | - | 5 | - | 3 | 1 | 2 | - | 10 | 2 | 3 | - | 1 | - | 1 | - |
| Ciprofloxacin (5 mcg) | 2 | 35 | 1 | 27 | - | 6 | - | 4 | - | 5 | - | 4 | - | 2 | 1 | 11 | - | 3 | - | 1 | - | 1 |
| Levofloxacin (5 mcg) | 2 | 35 | 2 | 26 | 1 | 5 | - | 4 | - | 5 | - | 4 | - | 2 | 1 | 11 | - | 3 | - | 1 | - | 1 |
| Cotrimoxazole (25 mcg) | 2 | 35 | 5 | 23 | 1 | 5 | 1 | 3 | 1 | 4 | - | 4 | - | 2 | 3 | 9 | - | 3 | 1 | - | - | 1 |

**Table 3.** Distribution spectrum of resistant and susceptible CRE (n=103) isolates among various bacterial spp against other commonly-used antimicrobial agents. *Klebsiella spp, Acinetobacter spp* showed maximum resistant isolated followed by *Pseudomonas spp, Escherichia coli spp, proteus spp*, and *Providencia spp*.

Sumon Das et. Al. 2022. Table\_3.
| Antimicrobial agents | Gram-negative antibiotic resistant organism |  |  |  |  |  |  |  |  |  |  |  |
| --- | --- | --- | --- | --- | --- | --- | --- | --- | --- | --- | --- | --- |
|  | *Escherichia coli* |  | *Klebsiella spp* |  | *Pseudomonas spp* |  | *Acinetobacter spp* |  | *Proteus spp* |  | *Providencia spp* |  |
|  | S | R | S | R | S | R | S | R | S | R | S | R |
| Amoxiclav (30 mcg) | - | 8 | - | 26 | - | 27 | - | 40 | - | 1 | - | 1 |
| Ceftriaxone (30 mcg) | - | 8 | - | 26 | - | 27 | 1 | 39 | - | 1 | - | 1 |
| Cefepime (30 mcg) | - | 8 | - | 26 | 4 | 23 | - | 40 | - | 1 | - | 1 |
| Ceftazidime (30 mcg) | - | 8 | - | 26 | 4 | 23 | 1 | 39 | - | 1 | - | 1 |
| Imipenem (10 mcg) | - | 8 | - | 26 | - | 27 | - | 40 | - | 1 | - | 1 |
| Meropenem (10 mcg) | - | 8 | - | 26 | - | 27 | - | 40 | - | 1 | - | 1 |
| Polymyxin (300 mcg) | 8 | - | 24 | 2 | 23 | 4 | 38 | 2 | 1 | - | - | 1 |
| Amikacin (30 mcg) | 4 | 4 | 1 | 25 | 5 | 22 | 1 | 39 | - | 1 | - | 1 |
| Gentamicin (10 mcg) | 3 | 5 | 4 | 22 | 2 | 25 | 2 | 38 | - | 1 | - | 1 |
| Netilmicin (30 mcg) | 2 | 6 | - | 26 | 4 | 23 | 3 | 37 | - | 1 | - | 1 |
| Tetracycline (30 mcg) | - | 8 | 10 | 16 | 7 | 20 | 2 | 38 | - | 1 | - | 1 |
| Tigecycline (15 mcg) | 8 | - | 24 | 2 | 12 | 15 | 38 | 2 | - | 1 | 1 | - |
| Ciprofloxacin (5 mcg) | - | 8 | 1 | 25 | 3 | 24 | - | 40 | - | 1 | - | 1 |
| Levofloxacin (5 mcg) | - | 8 | 1 | 25 | 3 | 24 | 2 | 38 | - | 1 | - | 1 |
| Cotrimoxazole (25 mcg) | 1 | 7 | 1 | 25 | 2 | 25 | 10 | 30 | - | 1 | - | 1 |

**Table 4.** Distribution spectrum of resistant bacterial CRE isolates (n=103) among different age-groups. against antimicrobial agents using the Kirby-Bauer disc diffusion techniques. Bacterial isolates from all age-group showed 100% resistance to Amoxiclav (30 μg), Imipenem (10 μg), and Meropenem (10 μg) while “61-75” age-group showed maximum number of resistant isolates.

| Antimicrobial agents | Age group |  |  |  |  |  |  |  |  |  |  |  |
| --- | --- | --- | --- | --- | --- | --- | --- | --- | --- | --- | --- | --- |
|  | 0-15 |  | 16-30 |  | 31-45 |  | 46-60 |  | 61-75 |  | 76->90 |  |
|  | S | R | S | R | S | R | S | R | S | R | S | R |
| Amoxiclav (30 mcg) | - | 5 | - | 7 | - | 13 | - | 27 | - | 34 | - | 17 |
| Ceftriaxone (30 mcg) | - | 5 | - | 7 | - | 13 | 1 | 26 | - | 34 | - | 17 |
| Cefepime (30 mcg) | - | 5 | - | 7 | 2 | 11 | - | 27 | 1 | 33 | 1 | 16 |
| Ceftazidime (30 mcg) | - | 5 | - | 7 | 2 | 11 | 1 | 26 | 1 | 33 | 1 | 16 |
| Imipenem (10 mcg) | - | 5 | - | 7 | - | 13 | - | 27 | - | 34 | - | 17 |
| Meropenem (10 mcg) | - | 5 | - | 7 | - | 13 | - | 27 | - | 34 | - | 17 |
| Polymyxin (300 mcg) | 5 | - | 6 | 1 | 13 | - | 25 | 2 | 29 | 5 | 16 | 1 |
| Amikacin (30 mcg) | - | 5 | 1 | 6 | 3 | 10 | 2 | 25 | 2 | 32 | 3 | 14 |
| Gentamicin (10 mcg) | - | 5 | 2 | 5 | 3 | 10 | 2 | 25 | 1 | 33 | 3 | 14 |
| Netilmicin (30 mcg) | 1 | 4 | 2 | 5 | 3 | 10 | 2 | 25 | 1 | 33 | - | 17 |
| Tetracycline (30 mcg) | 1 | 4 | 1 | 6 | 3 | 10 | 6 | 21 | 6 | 28 | 2 | 15 |
| Tigecycline (15 mcg) | 5 | - | 6 | 1 | 11 | 2 | 20 | 7 | 26 | 8 | 15 | 2 |
| Ciprofloxacin (5 mcg) | - | 5 | 1 | 6 | 3 | 10 | - | 27 | - | 34 | - | 17 |
| Levofloxacin (5 mcg) | - | 5 | 1 | 6 | 3 | 10 | 1 | 26 | - | 34 | 1 | 16 |
| Cotrimoxazole (25 mcg) | 2 | 3 | 1 | 6 | 3 | 10 | 4 | 23 | 1 | 33 | 3 | 14 |

**Figure 1.**
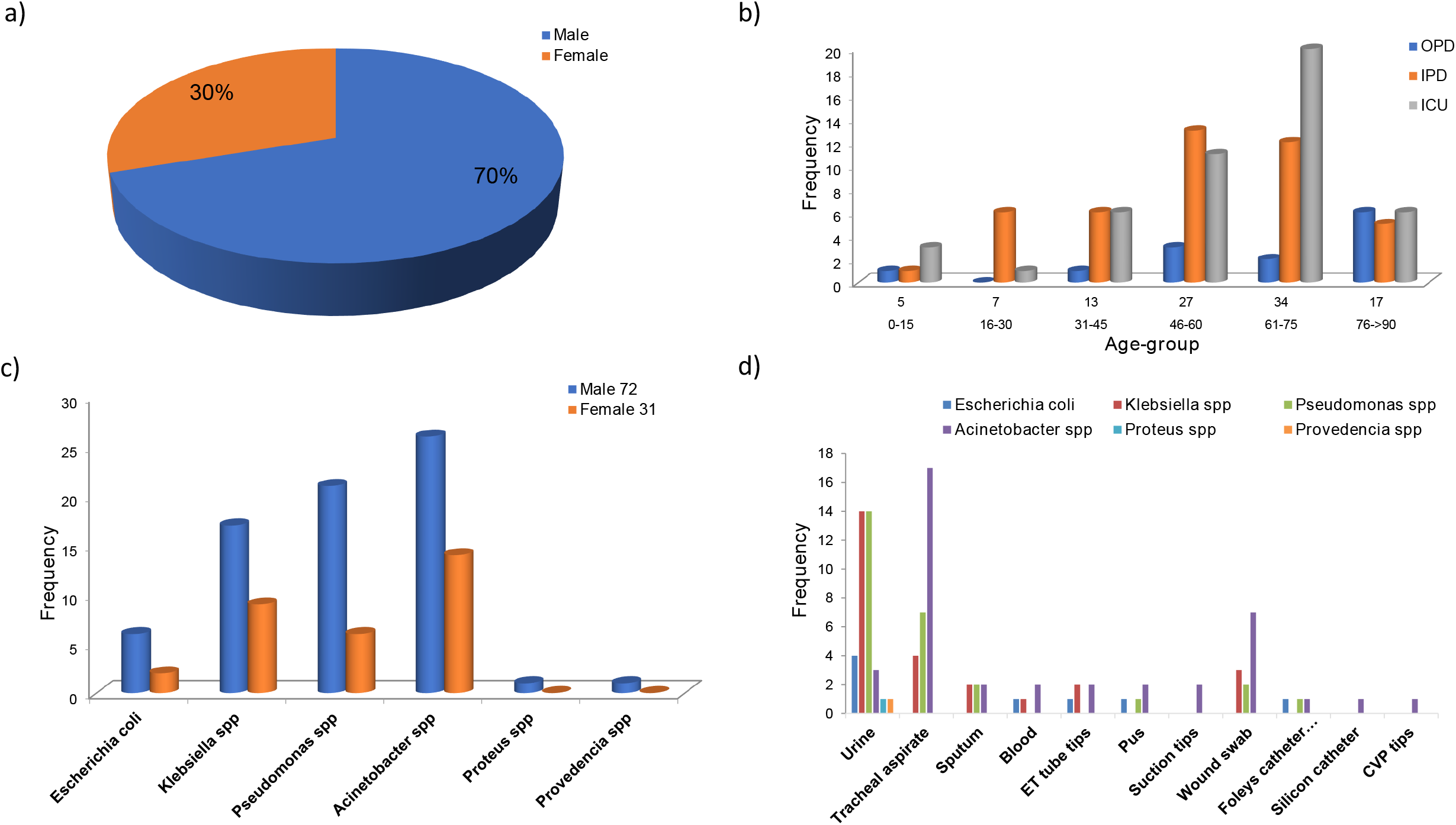
Bacterial isolates from all age-groups showed 100% resistance to Amoxiclav, and Carbapenems while “61-75” age-group showed maximum number of resistant isolates. (a) Showing the percentage of males and females affected by CRE. b). Frequency of clinically isolated etiological agent in clinical samples from various units in the hospital. c) Showing the frequency of CRE isolated according to age-group.

**Figure 2.**
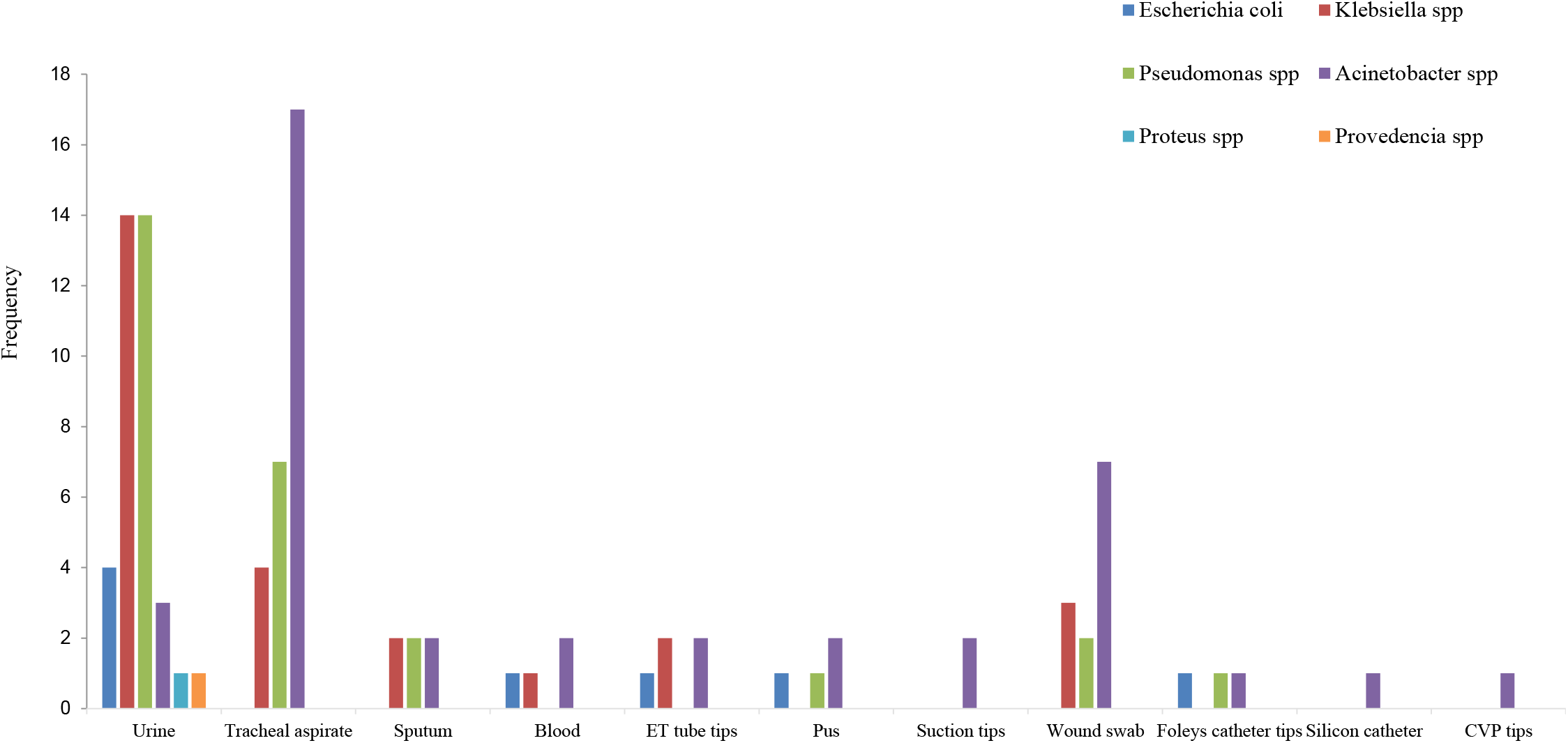
Sample-wise distribution spectrum of various gram-negative strains.

### The DPT testing method is more sensitive and detected higher number of MBL producers compared to the DDST

Among the five different DDPT and DDST methods were compared in this study, of 103 clinical isolates, 61 (59.2%) isolates showed sensitivity by disc potentiation test (DPT) method using imipenem with 0.1M EDTA and 56 (54.4%) isolates ceftazidime with 0.1M EDTA. In the DPT test method using imipenem with 0.1M EDTA, the highest resistance to carbapenem strains carrying MBL enzyme were found in IPD patient was 67% (29 out of 43) followed by ICU 53% (25 out of 47) and OPD 15% (07 out of 13) (Figure 3a). The inflated number of carbapenem resistant bacterial strains carrying MBL enzyme were found in age-group 0-15 years (80%, 4 out of 5) followed by 76->90 years (76%, 13 out of 17) (Figure 3b). The proportion of males infected with MBL enzyme-producing resistant bacteria was higher than the proportion of female (44 out of 72) and (17 out of 31) respectively (Figure 3c). The highest number of MBL-producing strain was *Escherichia coli* (8 out of 8) (Figure 3d). Using ceftazidime with 0.1M EDTA, the excessively high resistant strains to carbapenem carrying MBL enzyme were found in ICU patient (27 out of 47) followed by IPD (25 out of 43) and OPD (04 out of 13) (Figure 3a) and, the highest distribution were found in the age-group 61-75 years (19out of 34) (Figure 3b). The proportion of males infected with MBL enzyme-producing resistance bacterial strains was higher than the proportion of female (37 out of 72) and (19 out of 31) respectively (Figure 3c). The highest number of MBL-producing bacterial strain was *Klebsiella spp* (15 out of 26) followed by *Escherichia coli* (4 out of 8) (Figure 3d).

**Figure 3.**
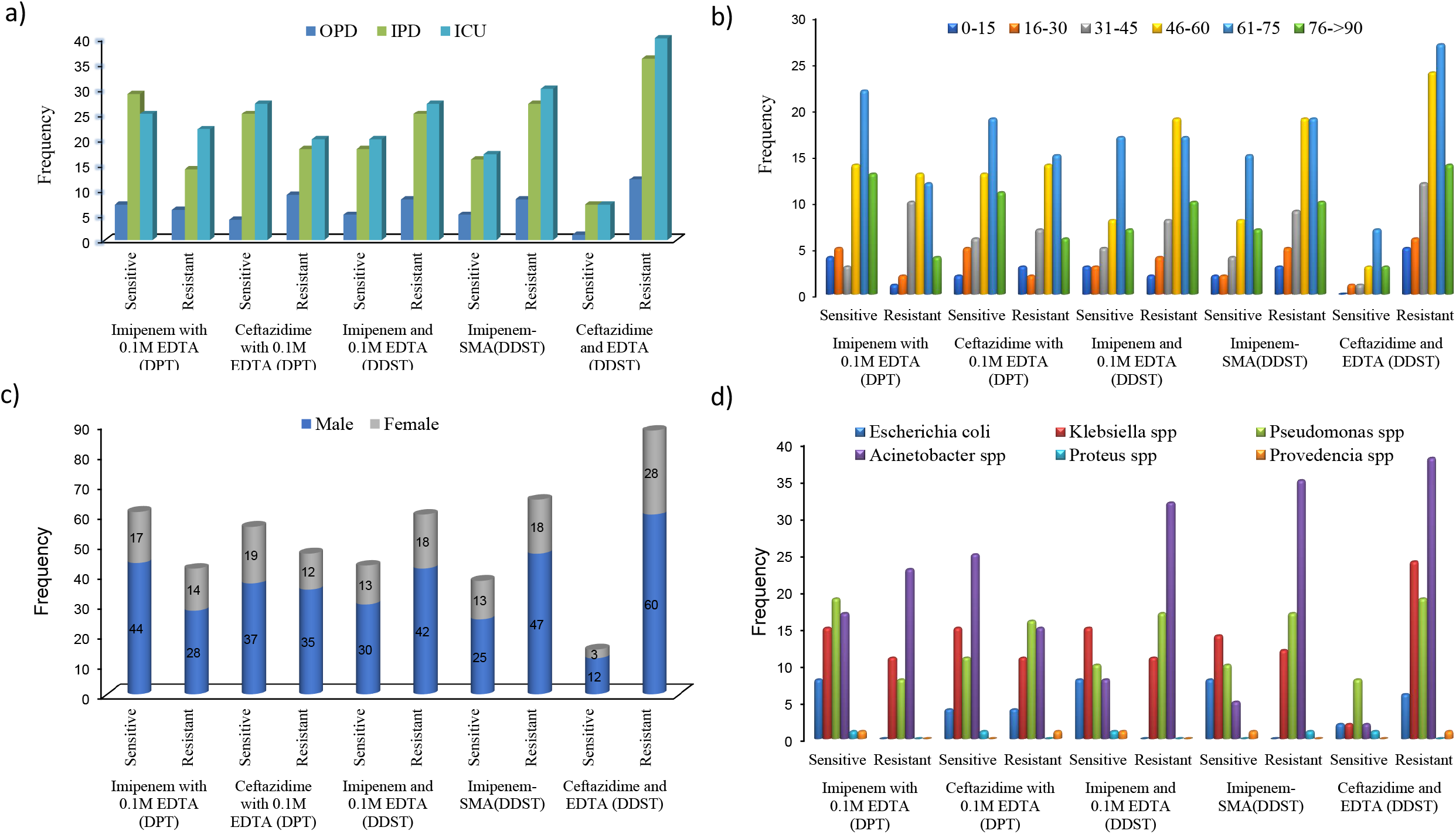
DPT testing method is more sensitive and detected higher number of MBL producers compared to the DDST. (a) Number of MBL enzyme screening using different inhibitor various unit of the hospital. (b) Showing the frequency of MBL enzyme isolated according to age-group (c) Number of MBL enzyme screening using different inhibitor affected by gender (d) Showing the frequency of MBL enzyme screening in different gram-negative strain.

In the DDST method using imipenem and 0.1M EDTA 43 (41.7%) isolates showed sensitive patterns followed by 38 (36.9%) imipenem and SMA and 15 (14.6%) isolates showed susceptibility to ceftazidime and EDTA. In this test method, using imipenem and 0.1M EDTA, the maximum resistant bacterial isolates to carbapenem strains carrying MBL enzyme were found in ICU patient (20 out of 47) followed by IPD (18 out of 43) and OPD (05 out of 13) (Figure 3a). The highest carbapenem-resistant bacterial isolates carrying MBL-enzyme were found in the age-group 61-75 years (17out of 34) (Figure 3b and the proportion of males infected with MBL enzyme-producing resistant bacterial strains was higher than the proportion of female (30 out of 72) and (13 out of 31) respectively (Figure 3c). The highest number of MBL-producing bacterial strain was *Escherichia coli* (8 out of 8) (Figure 3d). On the other hand, using imipenem and SMA, the highest number of bacterial isolates showing resistant to carbapenem carrying MBL-enzyme were found in ICU patient (17 out of 47) followed by IPD (16 out of 43) and OPD (05 out of 13) (Figure 3a), and the highest distribution of carbapenem resistant isolates carrying MBL-enzyme were found in age-group 61-75 years (15 out of 34) (Figure 3b) while the proportion of males patients infected with MBL enzyme-producing bacterial isolates was higher than of female (25 out of 72) and (13 out of 31) respectively (Figure 3c). The highest number of MBL producing bacterial strain was *Escherichia coli* (8 out of 8) (Figure 3d). Using ceftazidime and 0.1M EDTA, the highest number of bacterial isolates showing resistant to carbapenem strains MBL-enzyme was found in ICU patient (07 out of 47) followed by IPD (07 out of 43) and OPD (01 out of 13) (Figure 3a). The highest distribution of carbapenem resistant bacterial isolates carrying MBL-enzyme were observed in age-group 61-75 years (07 out of 34) and the proportion of males infected with MBL-enzyme producing resistant bacterial strains was higher than that of female (12 out of 72) and (03 out of 31) respectively (Figure 3b and 3c respectively), while the maximum number of MBL-producing bacterial strain was *Pseudomonas spp* (8 out of 27).

## Discussion

The global spread of carbapenemase-producing bacteria underlined the necessity of having tools available into all clinical diagnostic laboratories to monitor their emergence. Special attention needs to be given to the developing country like Bangladesh, where sheer negligence leads to a higher risk of spread. Not only negligence, availability of proper detection strategy of MBL-producing CRE leads to otherwise expensive detection system. Therefore, a cheaper, readily available and proper detection method for MBL producing CRE, especially in peripheral hospitals is necessary. Acquired MBLs in Gram-negative bacteria are becoming an emerging resistant determinant worldwide (11). To address this rising resistant determinant, we have investigated the prevalence and the distribution of MBL producers among the Imipenem-resistant isolates following standard methods of isolation and identification. Even though detection using PCR a simple and straightforward approach is gaining momentum worldwide (19), it comes with a considerable expense and usually not affordable and adaptable by most hospitals and clinics in Bangladesh. In contrast to molecular-genetic techniques, which are only able to detect genes that are already known, phenotypic detection of carbapenemase activity is also able to detect novel carbapenemases (1). Hence, two distinct processes DDST and DPT were interrogated for the best detection method to be identified, as these two processes are affordable and accessible to perform in most of the local hospitals.

Our study suggests, males are more susceptible compared to females to the resistant enterobacteriaceae no matter which location in Bangladesh they belong to, while the leading affected age-group was 61-75 years of age, most probable cause can be due to the weakened immune system at this age. A concerning issue was the highest percentage of bacterial identification in the ‘Ward unit’ of the Hospital. However, the total proportion of different ICU units exceeds the number of bacteria in the Ward unit (Figure 1.).

Reduced susceptibility of the MBL-producing Gram-negative bacteria to Imipenem has been frequently reported (20,21). Interestingly, many of these reports have documented only moderate resistance to Imipenem (21,22). The present study showed high levels (100%) of Imipenem and Meropenem resistance for all the Gram-negative bacteria tested (Table 1.). In addition, our analysis results on the multi-drug resistance (100%) represented that the highest number of resistant isolates were obtained from tracheal aspirate followed by urine and wound swabs however, were not affected by the sample types of the host (Figure 2. and Table 2.) origin. A significant increase in the percentage of *A. baumannii* isolates (38.8%) which causes infections in the blood, urinary tract, and lungs (pneumonia), or in wounds in other parts of the body resistant to Imipenem was observed in the present study (Table 3.) which is in contrast to the (15%) previously reported in the only other Bangladeshi study (11), reflecting the evolving scenario in Bangladesh. Our study demonstrated that that all MBL producers were of multidrug resistant. Selective pressure and/or the simultaneous presence of other drug resistance genes such as gene cassettes or other resistance mechanisms might be the reason for the co-resistance (11). The present study gave significant indications that DPT (combined disc test) using EDTA as an inhibitor and Imipenem as the substrate can be successfully used for the identification of MBL enzymes among the bacteria of the family *Enterobacteriaceae*. Although acquired MBL enzymes are frequently found in *Pseudomonas* spp. and *Acinetobacter* spp. however, this study suggests that the existence of MBL in the species of *E*.*coli* and *K*.*pneumoniae* might be due to plasmid-mediated horizontal transfer occurs continuously among Gram-negative bacilli, as reported previously (11).

The carbapenem-resistant strains with no MBL noticeable by the DPT test in this study may possess other enzymes mediating carbapenem resistance, such as OXA-type lactamases (class D) or AmpC b-lactamases, and/or other mechanisms such as outer-membrane permeability and efflux mechanisms (18) that were yet to check. Therefore, our research signifies that the identification of MBL-producing CRE by DPT as a better method compared to DDST (Figure 3.) in identifying the spread of multiple-antibiotic-resistant organisms. This finding will allow the development of containment measures, leading to broader impacts in reducing their transmission to the communities in Bangladesh. Moreover, our results highlight a handful of issues related to the prevalence and characteristics of MBL-producing Gram-negative bacteria in hospitals in Dhaka, a potential public health threat. Apart from the detection method, alarming fact about the rate of exposure of different age, sex groups, locations and specific sections in hospital was brought to light.

High incidence of MBLs-producers among carbapenem, and amoxiclav resistant Gram-negative micro-organisms obtained in this study, demands necessary implementations of strict infection control, rearranging the patient’s management including the detection methods to overcome the therapeutic challenges, reduce hospitalization-time constraint and treatment cost. Early detection of multi-drug resistant, implementation of strict antimicrobial policies and infection control programs by hospitals may avoid the rapid dissemination of these organisms. This investigation gives an apprehensive critique to dynamic detection of MBL-producing bacterial infection and will be of interest to the scientists, researchers, physicians working in hospital and microbiologists understanding, resistant mechanism, controlling infection, as well as to government policy makers to set up a guideline for managing hospitals.

## Data Availability

All relevant data are within the manuscript and its Supporting Information files.

## Acknowledgments

This work was partially supported by personal fund of Dr. N.Ara in Evercare Hospital Dhaka, Bangladesh.

All authors contributed to this research work from their responsibility to the society. We are grateful to the patients for providing samples and for their participation in our research.

## Legends

**Supplement Table 1.**
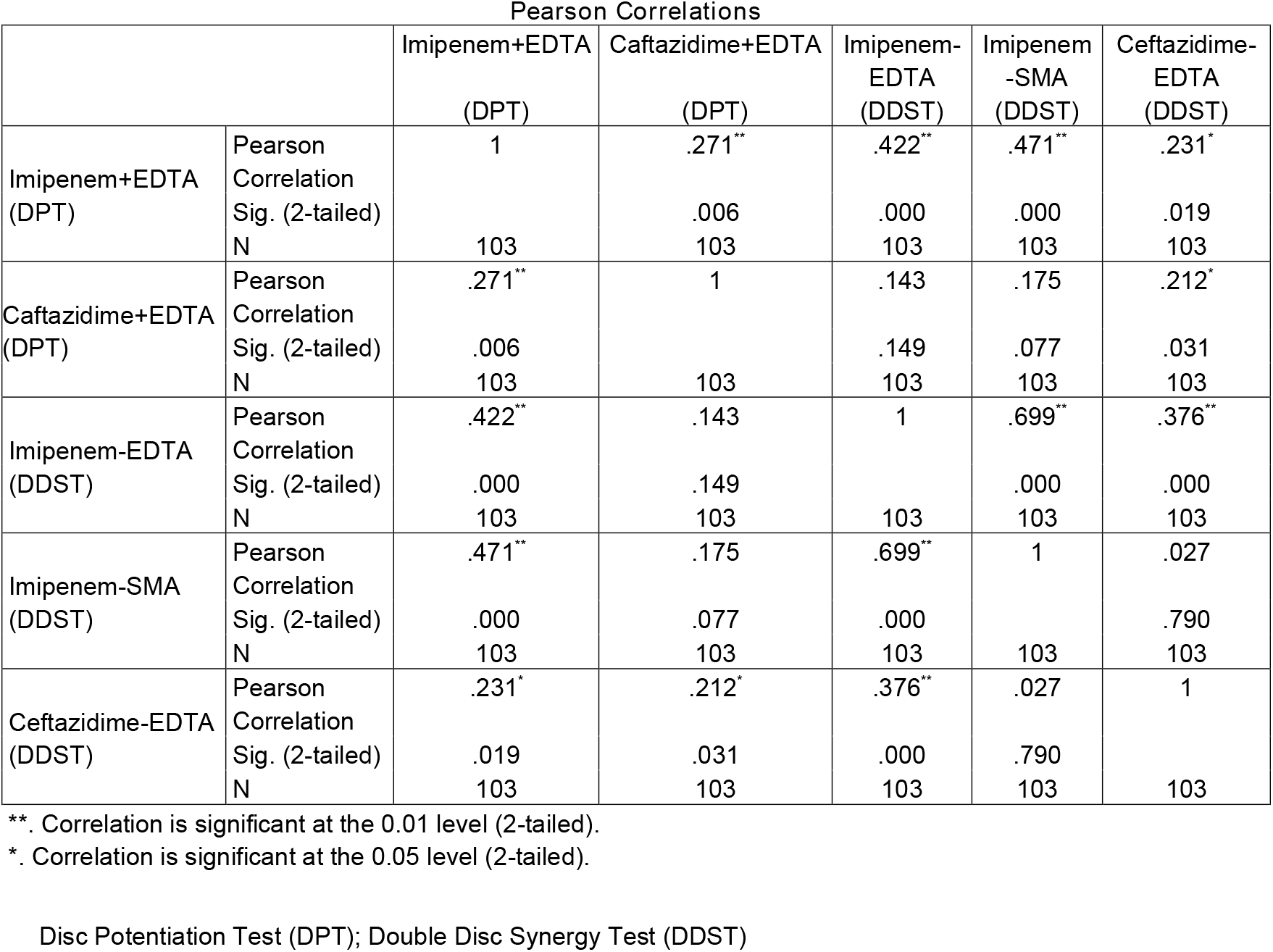
**Statistical significance of cross-Metallo-β-lactamases (MBLs), evaluated through the Disc Potentiation Test (DPT); Double Disc Synergy Test (DDST) were analyzed by the Pearson correlation test.**

## References

1. Hrabák J, Chudáčková E, Papagiannitsis CC. Detection of carbapenemases in Enterobacteriaceae: a challenge for diagnostic microbiological laboratories. Clinical Microbiology and Infection. 2014 Sep 1;20(9):839–53.

2. Kumarasamy KK, Toleman MA, Walsh TR, Bagaria J, Butt F, Balakrishnan R, Chaudhary U, Doumith M, Giske CG, Irfan S, Krishnan P. Emergence of a new antibiotic resistance mechanism in India, Pakistan, and the UK: a molecular, biological, and epidemiological study. The Lancet infectious diseases. 2010 Sep 1;10(9):597–602.

3. Andersson DI, Hughes D. Microbiological effects of sublethal levels of antibiotics. Nature Reviews Microbiology. 2014 Jul;12(7):465–78.

4. Islam MA, Islam M, Hasan R, Hossain MI, Nabi A, Rahman M, Goessens WH, Endtz HP, Boehm AB, Faruque SM. Environmental spread of New Delhi metallo-β-lactamase-1-producing multidrug-resistant bacteria in Dhaka, Bangladesh. Applied and environmental microbiology. 2017 Aug 1;83(15):e00793–17.

5. Tsakris A, Poulou A, Pournaras S, Voulgari E, Vrioni G, Themeli-Digalaki K, Petropoulou D, Sofianou D. A simple phenotypic method for the differentiation of metallo-β-lactamases and class A KPC carbapenemases in Enterobacteriaceae clinical isolates. Journal of antimicrobial chemotherapy. 2010 Aug 1;65(8):1664–71.

6. MerieQueenan A, Bush K. Carbapenemases: the versatile B-lactamases. Clin Microbiol Rev. 2007;20(3):440–58.

7. Naas T, Cuzon G, Villegas MV, Lartigue MF, Quinn JP, Nordmann P. Genetic structures at the origin of acquisition of the β-lactamase bla KPC gene. Antimicrobial agents and chemotherapy. 2008 Apr;52(4):1257–63.

8. Ikonomidis A, Tokatlidou D, Kristo I, Sofianou D, Tsakris A, Mantzana P, Pournaras S, Maniatis AN. Outbreaks in distinct regions due to a single Klebsiella pneumoniae clone carrying a bla VIM-1 metallo-β-lactamase gene. Journal of Clinical Microbiology. 2005 Oct;43(10):5344–7.

9. Picão RC, Cardoso JP, Campana EH, Nicoletti AG, Petrolini FV, Assis DM, Juliano L, Gales AC. The route of antimicrobial resistance from the hospital effluent to the environment: focus on the occurrence of KPC-producing Aeromonas spp. and Enterobacteriaceae in sewage. Diagnostic microbiology and infectious disease. 2013 May 1;76(1):80–5.

10. Goic-Barisic I, Hrenovic J, Kovacic A, Musić MŠ. Emergence of oxacillinases in environmental carbapenem-resistant Acinetobacter baumannii associated with clinical isolates. Microbial Drug Resistance. 2016 Oct 1;22(7):559–63.

11. Farzana R, Shamsuzzaman SM, Mamun KZ. Isolation and molecular characterization of New Delhi metallo-beta-lactamase-1 producing superbug in Bangladesh. The Journal of infection in developing countries. 2013 Mar 14;7(03):161–8.

12. Franklin C, Liolios L, Peleg AY. Phenotypic detection of carbapenem-susceptible metallo-β-lactamase-producing gram-negative bacilli in the clinical laboratory. Journal of clinical microbiology. 2006 Sep;44(9):3139–44.

13. Arakawa Y, Shibata N, Shibayama K, Kurokawa H, Yagi T, Fujiwara H, Goto M. Convenient test for screening metallo-β-lactamase-producing gram-negative bacteria by using thiol compounds. Journal of clinical microbiology. 2000 Jan 1;38(1):40–3.

14. Shibata N, Doi Y, Yamane K, Yagi T, Kurokawa H, Shibayama K, Kato H, Kai K, Arakawa Y. PCR typing of genetic determinants for metallo-β-lactamases and integrases carried by gram-negative bacteria isolated in Japan, with focus on the class 3 integron. Journal of Clinical Microbiology. 2003 Dec;41(12):5407–13.

15. Notake S, Matsuda M, Tamai K, Yanagisawa H, Hiramatsu K, Kikuchi K. Detection of IMP metallo-β-lactamase in carbapenem-nonsusceptible Enterobacteriaceae and non-glucose-fermenting Gram-negative rods by immunochromatography assay. Journal of clinical microbiology. 2013 Jun;51(6):1762–8.

16. Yong D, Lee K, Yum JH, Shin HB, Rossolini GM, Chong Y. Imipenem-EDTA disk method for differentiation of metallo-β-lactamase-producing clinical isolates of Pseudomonas spp. and Acinetobacter spp. Journal of clinical microbiology. 2002 Oct;40(10):3798–801.

17. Galani I, Rekatsina PD, Hatzaki D, Plachouras D, Souli M, Giamarellou H. Evaluation of different laboratory tests for the detection of metallo-β-lactamase production in Enterobacteriaceae. Journal of antimicrobial chemotherapy. 2008 Mar 1;61(3):548–53.

18. Karthika RU, Rao RS, Sahoo S, Shashikala P, Kanungo R, Jayachandran S, Prashanth K. Phenotypic and genotypic assays for detecting the prevalence of metallo-β-lactamases in clinical isolates of Acinetobacter baumannii from a South Indian tertiary care hospital. Journal of medical microbiology. 2009 Apr 1;58(4):430–5.

19. Walsh TR, Toleman MA, Poirel L, Nordmann P. Metallo-β-lactamases: the quiet before the storm?. Clinical microbiology reviews. 2005 Apr;18(2):306–25.

20. Joshi SG, Litake GM, Ghole VS, Niphadkar KB. Plasmid-borne extended-spectrum β-lactamase in a clinical isolate of Acinetobacter baumannii. Journal of medical microbiology. 2003 Dec 1;52(12):1125–7.

21. Sinha M, Srinivasa H. Mechanisms of resistance to carbapenems in meropenem-resistant Acinetobacter isolates from clinical samples. Indian journal of medical microbiology. 2007 Apr 1;25(2):121–5.

22. Taneja N, Sharma M. Imipenem resistance in nonfermenters causing nosocomial urinary tract infections. Indian journal of medical sciences. 2003 Jul 1;57(7):NA-.

23. Usman J, Kaleem F, Khalid A, Iqbal M. Detection and antibiotic susceptibility pattern of biofilm producing Gram positive and Gram.

